# How Many Lives Has Lockdown Saved in the UK?

**DOI:** 10.1101/2020.06.24.20139196

**Authors:** Rickard Nyman, Paul Ormerod

## Abstract

We compare the trajectory of deaths (both in hospitals and care homes) on a daily basis in Sweden and England and Wales (which constitute 90 per cent of the UK population) from 11 March to 7 August 2020, the latest date at which the relevant data is available for England and Wales.

Deaths in both Sweden and England and Wales peaked on 8 April. The build up to the peak was very similar in both. Given the time lag between infection and death, the lockdown would have had little effect on the peak number of deaths.

By the first week of August, the deaths are very similar in both. However, from early May the decline in England and Wales has been much sharper.

We estimate that to 7 August, lockdown saved 17,700 lives in England and Wales, or just under 20,000 extrapolating to a UK level.

## 1. Introduction and Background

Professor Ferguson and the Imperial College team, in a highly influential paper of 16 March 2020^3^, argued that without any policy changes “we predict approximately 510,000 deaths in Great Britain” (p.7). They also suggested that the peak of mortality would take place after approximately 3 months.

In this paper, we compare the trajectories of Covid deaths in the UK and Sweden from early March to early August in order to form an assessment of how many lives have been saved in the UK by lockdown.

In the UK, an “indicative” regime was introduced by the Prime Minister in a broadcast to the nation on the evening of 16 March. It was left to individuals to decide how to behave. Very shortly afterwards, it was followed by a rather strict list of legally based measures, announced on the evening of 23 March.

Sweden, in comparison to other Western countries, has notoriously operated a much more relaxed policy towards lockdown. There have been some legal restrictions in Sweden^4^, but they have been much lighter than those applied in the UK. For example, public (but not private) gatherings of more than 50 people have been banned. However, bars and restaurants have remained open with few restrictions.

Sweden is therefore a useful comparator with which to assess the experience of the UK.

We note initially that dire predictions of mortality in Sweden were made in the earlier stages of the pandemic. For example, using a model applied to Sweden based on the Imperial one, on 15 April 2020, Gardner and colleagues^5^ stated that the model predictions indicated that “at least 96,000 deaths would occur by 1 July without mitigation. Current policies reduce this number by approximately 15%” (p.4).

Covid-related deaths in Sweden have been *far* lower than the predictions suggested. Taking the daily death data from https://www.folkhalsomyndigheten.se/, the Public Health Agency of Sweden, the total number of deaths over the period 11 March to 7 August 2020 is only 5776 (this includes deaths in care homes).

Of course, we do not yet know whether there will be a major second wave of the virus later in the summer, something which models such as the Imperial one are only too ready to predict. For example, Ferguson and his colleagues at Imperial College emphasised on 30 March “the virus will be able to spread rapidly should interventions be lifted” (p.12)^6^. The same group published projections for Italy on 4 May 2020^7^. They concluded that even modest returns towards the pre-lockdown levels of social mobility “may lead to a resurgence of the epidemic with more deaths than the current wave in the absence of additional interventions” (p.17).

But there is a huge difference between the Swedish projections and the outcome so far. The predictions were for just over 80,000 deaths. By 7 August, the actual figure to the middle of June is only just under 6000..

The Office for National Statistics in the UK publishes death data on a daily basis for England and Wales. This include all deaths both inside and outside hospitals, including care homes (which are also included in the Swedish data). England and Wales make up 90 per cent of the population of the UK as a whole.

## 2. Comparing the time paths of deaths from Covid

### 2.1 Similarities

There are three important similarities between the two series over the period 11 March to 7 August (respectively, the date on which the Swedish data starts and the latest available date for EW).

First, at the start of the data on deaths, in total on 11 and 12 March Sweden reported a single death. Adjusting the England and Wales (EW for short) data for the respective populations (10.23 million in Sweden, 59.88 million in England and Wales) gives a figure of 2.9 deaths in total on those days. In the week to 7 August, there were 16 deaths in Sweden and 18 in England and Wales (population adjusted to that of Sweden). The two series therefore start and finish at very similar levels.

Second, in both countries, the peak number occurs on 8 April (though the identical number of deaths to 8 April was also observed in Sweden on 15 April).

The 8 April peak in deaths corresponds to a peak in new cases in the week 18-25 March, given that the typical time from infection to death is in the range 14-21 days^8^. The voluntary lockdown was introduced in EW on the evening of 16 March and the compulsory one on the evening of 23 March. So they were in effective operation on 17 and 24 March. We can reasonably suppose that these had little influence in determining the peak level of infections and hence the peak level of deaths in early April.

A useful way of comparing the build up to the peak level is to examine the dates on which deaths reached both a quarter and a half of the eventual peak. This reveals the third similarity between the series.

The “quarter peak” number of deaths was reached in Sweden on 26/27 March and in EW on 25/26 March. The “half peak” was on 1/2 April in Sweden and 30/31 March in EW.

### 2.2 Differences

There are, however, two important differences between the two series, the second of which will be the basis of our estimate of the number of lives saved by lockdown in the UK. These are both illustrated in Figure 1 below.

**Figure 1.**
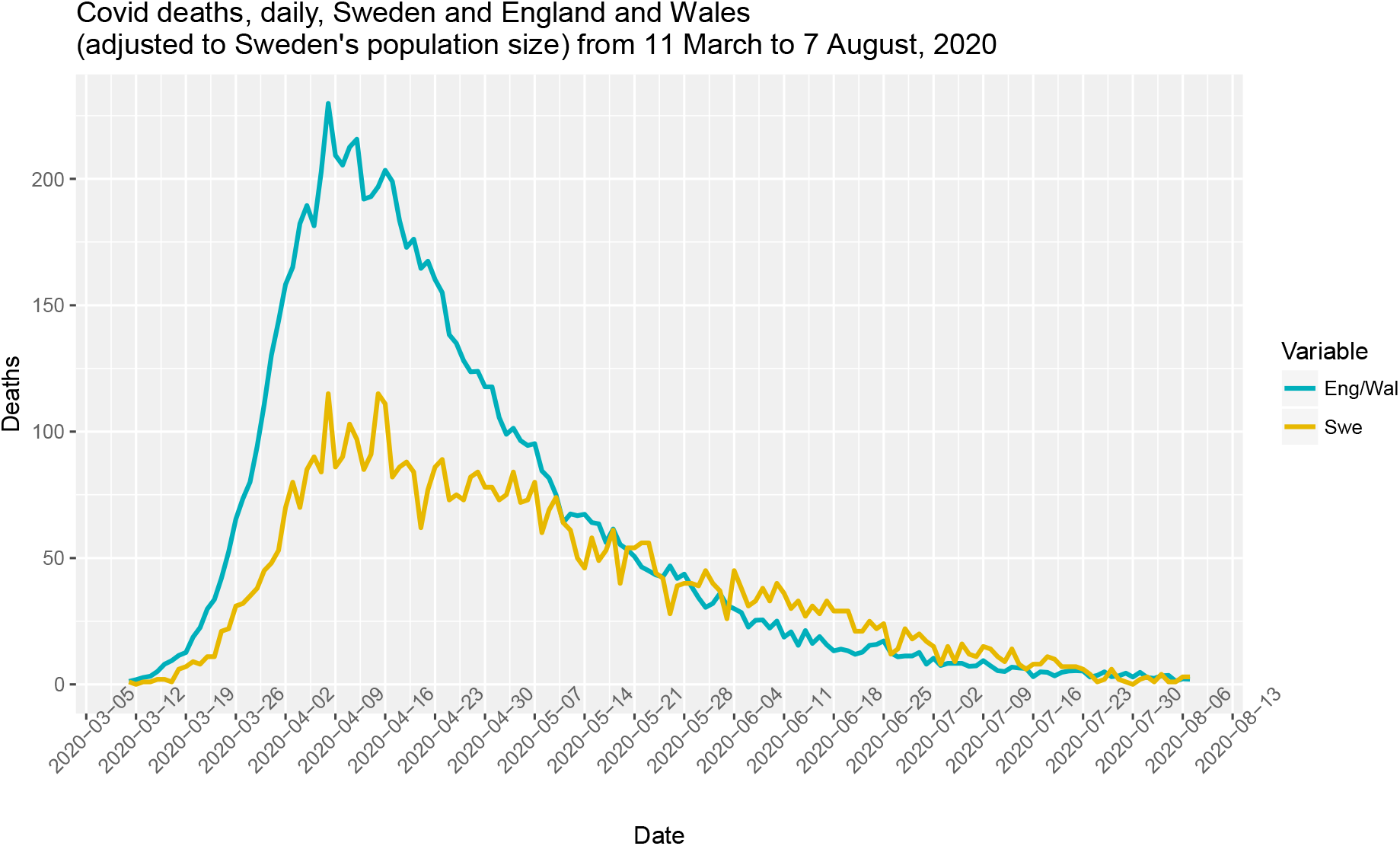
Daily total of deaths in Sweden and England and Wales, the latter adjusted to the Swedish population level. Sources: Folkhalsomyndigheten and Office of National Statistics

The first difference is that, adjusting the data for the relative sizes of the populations, the peak value in EW was almost exactly double that of Sweden.

It is not our purpose to offer a detailed explanation of this. As we argued above, the build up to the peak was, at best, only marginally influenced by the lockdowns.

However, two potential reasons spring to mind readily. First, obesity has been identified as an important determinant of deaths, and the UK rate of 27.8 per cent is higher than Sweden’s 20.6 per cent (https://www.cia.gov/library/publications/the-world-factbook/rankorder/2228rank.html). Second, EW has a much higher population density than Sweden and there is good evidence that density is a key factor in transmission of Covid^9^.

The second difference is that the pattern of decline of deaths from the peak is different in Sweden and EW.

The decline patterns are different. EW dropped to “half peak” by 30 April/1 May and “quarter peak” by 17/18 May.

Taking the first Swedish peak as the peak, “half peak” was not until around 11/12 May and “quarter peak” was not really until 14/15 June (there is a single day, 26 May, at quarter-level, but the observations either side are around one-third peak).

As noted above, the peak level in EW is twice that of Sweden, but the number of deaths falls away more sharply in EW.

The trajectory of the decline after the peak is shown more clearly in Figure 2, which also illustrates the very similar nature of the rises to the peak. This plots the number of deaths, with, in each country, the data indexed so that the peak level (on 8 April in each case) is equal to 100.

**Figure 2.**
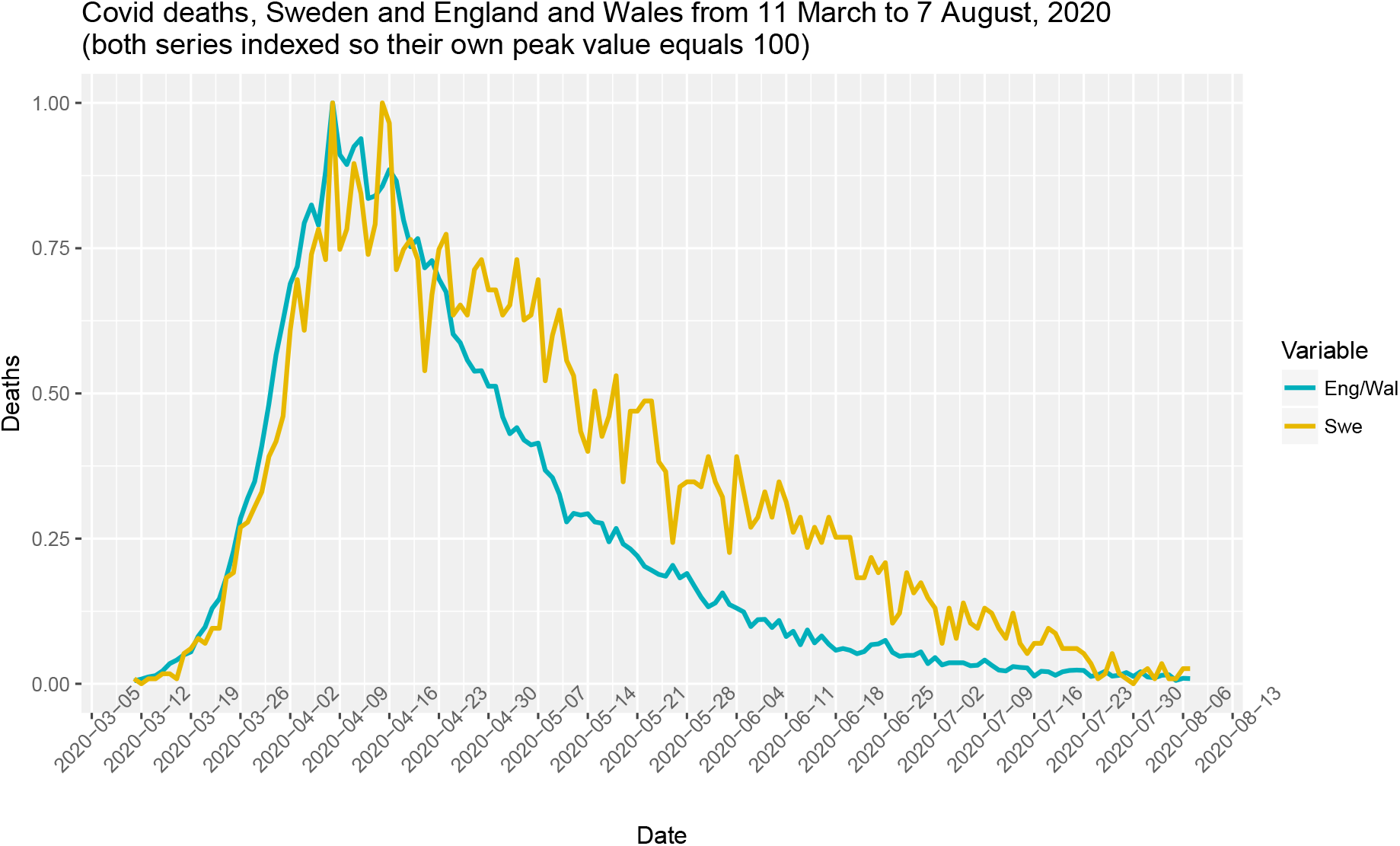
Daily total of deaths in Sweden and England and Wales, in each case adjusted so that the peak level (on 8 April) =100

We can use Figure 2 to form an estimate of the number of lives saved by the more rigorous lockdown in EW.

We calculate the cumulative value of the index in each country from 8 April, the peak value, onwards. There are some initial fluctuations in both countries, but by 29 April the cumulative value from 8 April is equalised in the two countries.

From 30 April until 7 August, the cumulative value of the index is distinctly higher in Sweden than it is in EW. In terms of index points, the difference is 13.16. In the EW data, an index value of 1 is equivalent to 1349 deaths, the number which occurred in EW on 8 April. We therefore estimate that the number of lives saved in EW by the lockdown is 1349 multiplied by 13.16, which is 17,751.

## 3. Brief discussion

Compared to the dramatic predictions about death levels if lockdown were not imposed, this number is very low. However, many of the standard models used in epidemiology do not take account of the fact that, faced by a pandemic, people alter their behaviour of their own accord. They do not wait for the decrees of the authorities.

World history is replete with examples. The Black Death in the 14^th^ century seems to have been the most deadly virus in recorded history. As much as 50 per cent of the population of Britain may have died. People adopted many different strategies to try and avoid the disease, some successful, others not.^10^ Giving lockdown a legal basis merely reinforces the behavioural adjustments which many people make when faced with a pandemic.

We note, finally, that a larger proportion of deaths took place in care homes in Sweden than in England and Wales. It could therefore be argued that even our estimate of 17,700 is perhaps on the high side.

## Data Availability

data for England and Wales available from https://www.ons.gov.uk/peoplepopulationandcommunity/birthsdeathsandmarriages/deaths/bulletins/deathsregisteredweeklyinenglandandwalesprovisional/weekending12june2020 and data for Sweden available from https://www.folkhalsomyndigheten.se/smittskydd-beredskap/utbrott/aktuella-utbrott/covid-19/bekraftade-fall-i-sverige/

https://www.ons.gov.uk/peoplepopulationandcommunity/birthsdeathsandmarriages/deaths/bulletins/deathsregisteredweeklyinenglandandwalesprovisional/weekending12june2020

## Footnotes

3 N Ferguson et al., 16 March 2020, Report 9: Impact of non-pharmaceutical interventions (NPIs) to reduce COVID-19 mortality and healthcare demand, DOI: https://doi.org/10.25561/77482

4 See https://polisen.se/aktuellt/nyheter/2020/mars/ytterligare-begransade-mojligheter-till-allmanna-sammankomster-och-tillstallningar

5 JM Gardiner et al., 15 April 2020, Intervention strategies against COVID-19 and their estimated impact on Swedish healthcare capacity, medRxiv, doi: https://doi.org/10.1101/2020.04.11.20062133

6 Imperial College COVID-19 Response Team DOI: https://doi.org/10.25561/77731, 30 March 2020, Estimating the number of infections and the impact of non-pharmaceutical interventions on COVID-19 in 11 European countries

7 N Ferguson et al., 4 May 2020, Using mobility to estimate the transmission intensity of COVID-19 in Italy: A subnational analysis with future scenarios, DOI: https://doi.org/10.25561/78677

8 For example, N Ferguson et al. Imperial College COVID-19 Response Team DOI: https://doi.org/10.25561/77731, 30 March 2020, Estimating the number of infections and the impact of non-pharmaceutical interventions on COVID-19 in 11 European countries

9 B Rader et al., 20 April 2020, Crowding and the epidemic intensity of COVID-19 transmission, medRxiv preprint doi: https://doi.org/10.1101/2020.04.15.20064980, version posted 20 April.

10 The excellent book by Ben Gummer describes this in detail. The Scourging Angel: The Black Death in the British Isles, Vintage, 2010

## Notes

### Competing Interest Statement

The authors have declared no competing interest.

### Funding Statement

No external funding was received for this work

### Author Declarations

Not applicable as the study is based entirely on an analysis of publicly available data

### Summary of Updates

Updated previous estimates to incorporate data from the past 2 months and revisions to previous data.

